# Classification of schizophrenia spectrum disorder using machine learning and functional connectivity: reconsidering the clinical application

**DOI:** 10.1101/2020.05.30.20118026

**Authors:** Chao Li, Fei Wang, Xiaowei Jiang, Ji Chen, Jia Duan, Shaoqiang Han, Hao Yan, Yanqing Tang, Ke Xu

## Abstract

An accurate identification of schizophrenia spectrum disorder (SSD) at early stage could potentially allow for treating SSD with appropriate intervention to potentially prevent future deterioration. Despite mounting studies found neuroimaging combined with machine learning can identify chronic medicated SSD, whether or not the classification model identified the trait biomarker of SSD that can be used to identify early stage SSD is largely unknown. The present study aimed to investigate whether or not the classification model trained using chronic medicated SSD identified the trait biomarker of SSD that whether or not the model can be generalized to early stage SSD, by using functional connectivity (FC) combined with support vector machine (SVM) using a large sample from 4 independent sites (n = 1077). We found that the classification model trained using chronic medicated SSD from three sites(dataset 2, 3 and 4) classified SSD from HCs in another site (dataset 1) with 69% accuracy (P = 2.86e-13). Subgroup analysis indicated that this model can identify chronic medicated SSD in dataset 1 with 71% sensitivity (P = 4.63e-05), but cannot be generalized to first episode unmedicated SSD (sensitivity = 48%, P = 0.68) and first episode medicated SSD (sensitivity = 59%, P = 0.10). Univariable analysis showed that medication usage had significant effect on FC, but disease duration had no significant effect on FC. These findings suggest that the classification model trained using chronic medicated SSD may mainly identified the pattern of chronic medication usage state, rather than the trait biomarker of SSD. Therefore, we should reconsider the current machine learning studies in chronic medicated SSD more cautiously in term of the clinical application.

## Introduction

Growing evidence from researches and clinical experiences demonstrated that signs and symptoms of psychiatric disorders do not map well to their neurobiological abnormalities (1). As one of the most 2 / 19 serious psychiatric disorder, schizophrenia, at present, however, its diagnosis, subtyping, prognosis, and treatment selection is mainly based on signs and symptoms, without clinically usable biomarkers to help doctors make clinical decisions, e.g., early diagnosis or prediction of disease onset. A much less optimistic situation is that antipsychotic medications are not effective in about one-third of patients (2,3). The gap between neurobiological endophenotypes and symptomatic phenotypes as well as the heterogeneous (4, 5) may largely contribute to the missed diagnosis and misdiagnosis, which further hinder the effective treatment. Undoubtedly, it is an urge to build and valid a clinically usable objective biomarker to early diagnose schizophrenia.

In recent years, there has been an increasing interest in the use of neuroimaging combined with machine learning for the diagnosis of schizophrenia spectrum disorder (SSD). Mounting studies have shown that functional and structural MR imaging can discriminate patients with SSD from healthy populations in the individual level (6–10). Previous studies using SVM and other machine learning methods found that unimodal or multimodal MR imaging could achieve 72%-80% accuracy to differentiate SSD from healthy controls (HCs) (11–17). For example, a recent study reported promising results that function and connectivity of the striatum may be a potential trait marker for schizophrenia using functional MR imaging dataset from seven independent centers in China (20). On the other hand, another prior study (21) found that structural neuroimaging cannot diagnose the first episode psychosis using 5 independent datasets.

Although growing vigorously researches has been carried out, there is still some key questions need to be answered (22–24). First, can we repeat the good diagnostic performance of studies with small sample size by using multi-center datasets, so as to confirm the reliability and generalization? Second, considering that most of the patients in the previous studies were chronic and medicated, can the trained model be generalized to the early stage patients or just limited in chronic medicated SSD? The second is crucial because, in clinic, it is very important to identify a trait biomarker to diagnose the psychiatric patients who are at the early stage and treatment plan are uncertain. If disease could be identified early on, appropriate intervention may potentially prevent future deterioration of this disease (25, 26).

The aim of this study was to investigate whether or not the trained classification model using samples of chronic medicated SSD can be generalized to early stage SSD by using FC combined with SVM. To avoid over-fitting problem, in the large sample study, we trained a classification model in three independent sites, and then test the model in an unseen dataset in another site.

## Methods

### Participants

We included four datasets from four centers respectively, which includes 1077 participants (502 patients with SSD and 575 HCs). All participants provided written informed consent, according to the procedures approved by the Ethics Committee or Institutional Review Boards of the respective hospitals or centers. The four centers were The First Affiliated Hospital of China Medical University (dataset 1: 188 patients with schizophrenia, 87 patients with schizophreniform psychosis and 275 HCs; among which were 92 chronic medicated SSD, 74 first episode medicated SSD and 44 first episode unmedicated SSD, Figure S2 showed their demographic and clinical information), Peking University Sixth Hospital (dataset 2: 106 patients with schizophrenia and 100 HCs), Center for Biomedical Research Excellence (dataset 3: 71 patients with schizophrenia and 74 HCs; data available at (http://fcon_1000.projects.nitrc.org/indi/retro/cobre.html) and University of California, Los Angles (dataset 4: 50 patients with schizophrenia and 126 HCs; data available at OpenfMRI database [accession number 4 / 19 is ds000030]). All patients were evaluated by qualified psychiatrists and diagnosed as schizophreniform psychosis or schizophrenia using the Structured Clinical Interview for DSM-IV-TR Axis I Disorders (SCID-I, patient edition). For participants younger than 18 years in the dataset 1, the Schedule for Affective Disorders and Schizophrenia for School-Age Children–Present and Lifetime Version (K-SADS-PL) was used for diagnosis. No patients had a history of neurological disorder, history of mental retardation, history of severe head trauma, history of substance abuse or electroconvulsive therapy. Please refer to the follow Supplementary Materials for the detailed inclusion and exclusion criteria of patients and HCs for dataset 1 and dataset 2. Please refer to the follow website for the detailed inclusion and exclusion criteria for dataset 3 and dataset 4 respectively: http://fcon_1000.projects.nitrc.org/indi/retro/cobre.html and https://www.openfmri.org/dataset/ds000030/. The symptoms severity of the patients was measured with the Brief Psychiatric Rating Scale (BPRS) in dataset 1 and Positive and Negative Syndrome Scale (PANSS) in dataset 2. The demographic and clinical characteristics are presented in Table 1. Here we defined the first episode SSD as having illness duration equal or less than 18 months and defined the chronic SSD as having illness duration more than 18 months. At present, there is no consistent result to distinguish the long and short duration of SSD, so we additionally used 24-month and 36-month thresholds to define first episode and chronic SSD (Supplementary Materials Figure S5).

| Sites | Group | Age (y) | Sex<br>(M/F) | BPRS | Positive<br>scale | Negative<br>scale | General<br>scale | Duration<br>(months) |
| --- | --- | --- | --- | --- | --- | --- | --- | --- |
| First Affiliated<br>Hospital of China<br>Medical<br>University (all) | Patient<br>(n = 275) | 25.0 ± 9.8 | 109/166 | 31.8 ± 12.4<br>(n = 267) | — | — | — | 32.9 ± 50.9<br>(n =244) |
|  | Control<br>(n = 275) | 25.3 ± 6.5 | 113/162 | 18.6 ± 1.9<br>(n = 268) | — | — | — | — |
| First Affiliated<br>Hospital of China<br>Medical<br>University (first<br>episode<br>unmedicated) | Patient<br>(n = 44) | 24.0 ± 9.8 | 26/18 | 41.2 ± 12.3<br>(n = 40) | — | — | — | 3.8 ± 4.1 |
|  | Control<br>(n = 44) | 24.4 ± 7.35 | 28/16 | 18.6 ± 1.3<br>(n = 40) | — | — | — | — |
| Peking University<br>Sixth Hospital | Patient<br>(106) | 27.1 ± 6.7 | 43/63 | — | 25.8 ± 5.4 | 21.5 ± 6.9 | 37.7 ± 6.2 | 53.0 ± 52.9 |
|  | Control<br>(100) | 25.8 ± 5.4 | 47/53 | — | — | — | — | — |
| COBRE | Patient<br>(71) | 38.1 ± 14.0 | 57/14 | — | — | — | — | 192.5 ±<br>149.9<br>(n = 70) |
|  | Control<br>(74) | 35.8 ± 11.6 | 51/23 | — | — | — | — | — |
| UCLA | Patient<br>(50) | 36.5 ± 8.9 | 38/12 | — | — | — | — | — |
|  | Control<br>(126) | 31.12 ± 8.7 | 59/67 | — | — | — | — | — |

### Image acquisition

All participants underwent scanning for 5–8 minutes using 3.0 Tesla MRI equipments. The detailed imaging acquisition protocols for the four datasets are provided in the Supplementary Materials.

### Overview of Methodology

Figure 1 presents an overall flowchart of the study. First, the standard preprocessing procedure was applied to these raw datasets of the 4 centers, followed by whole-brain ROI-wise FC network constructing by using Pearson’s correlation analysis. We inspected the average FC patterns of the four centers and their correlation, making sure that the data variance was acceptable (Figure 2). Then, we trained an SVM classifier using functional connectivity in three datasets: dataset 2, dataset 3 and dataset 4 (227 SSD, 300 healthy controls [HCs]; SSD in these datasets were all chronic medicated). Then, we used this trained classification model to predict the unseen samples in dataset 1 (275 SSD, 275 HCs). Because dataset 1 contained chronic medicated SSD (N = 92), first episode medicated SSD (N = 74) and first episode unmedicated SSD (N = 44), so we can investigate classification performance of the model on these 3 subgroups of SSD. Besides, we also tried other machine learning strategies: 5-fold cross-validation that pooled all datasets, five-fold cross-validation that only including first episode unmedicated SSD and leave-one-site-out cross-validation (please see the Supplementary Materials for details). All analyses codes are available here: https://github.com/lichao312214129/lc_rsfmri_tools_python/tree/master/Workstation/SSD_classification

**Figure 1.**
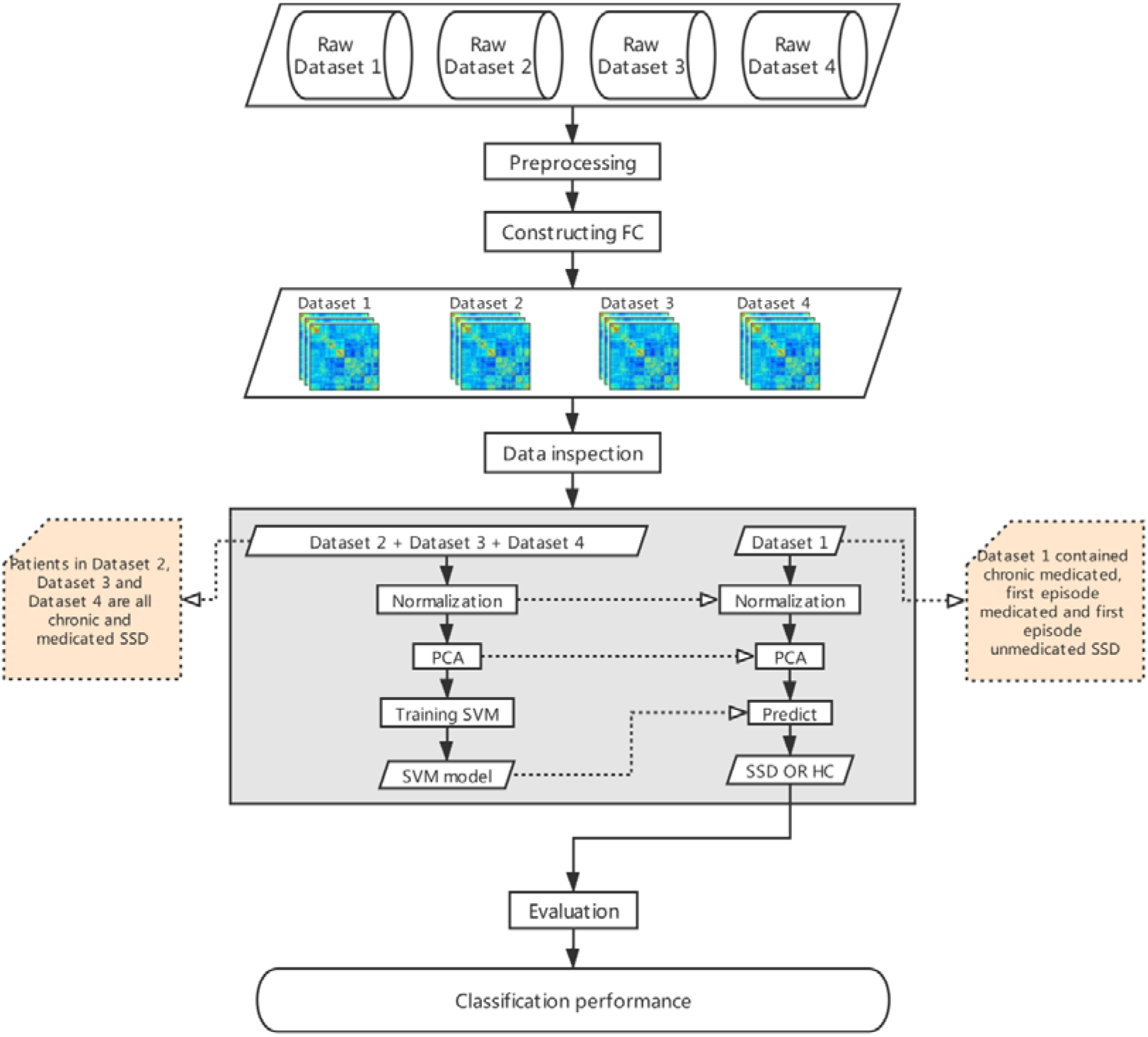
Flowchart of the study. We collected resting-state functional MRI data from four centers. The standard preprocessing procedure was applied to these data, followed by whole-brain ROI-wise FC network constructing. Then, we trained a SVM classification model using dataset 2, dataset 3 and dataset 4. Then, we used this trained classification to predict the unseen samples in dataset 1. Because dataset 1 contained chronic medicated SSD, first episode medicated SSD and first episode unmedicated SSD, so we can also investigate classification performance of the trained model on these 3 subgroups of SSD. SSD = Schizophrenia Spectrum Disorder; HC = Healthy Control; PCA = Principal Component Analysis; SVM = Machine.

### Data preprocessing

All images were preprocessed using SPM12 (http://www.fil.ion.ucl.ac.uk/spm/) and Data Processing & Analysis of Brain Imaging (DPABI) (27). The volumes from the first 10 time points were discarded. The subsequent preprocessing steps included slice-timing correction, head motion correction, spatially normalization (EPI template in Montreal Neurological Institute space, resampled to 3 mm × 3 mm × 3 mm isotropic voxels), smoothing (Gaussian kernel with a 4 mm full width at half-maximum), linear detrending, temporal bandpass filtering (0.01–0.01 Hz) and confounding covariates regressing (including the Friston-24 head motion parameters, white matter, cerebrospinal fluid, and global signals).

### FC network construction

The average time series of 246 nodes within the Brainnetome atlas (28) were extracted for each individual by averaging the whole time series throughout all voxels in each node. FC between each pair of nodes was calculated using Pearson’s correlation analysis, producing (246×245)/2 = 30135 unique FCs for each subject. Fisher r-to-z transformation was performed for all FCs.

### Machine learning

We pooled the dataset 2, dataset 3 and dataset 4 as the training data and used the dataset 1as the test data. The z normalization (divide by mean and subtract standard deviation) was applied to normalize data. We used principal component analysis (PCA) to reduce feature dimensionality. The top principal components with the highest eigenvalues that cumulatively explained 95% of the variance were selected (70%, 80% and 99% explained variance were also tried, please see Supplementary Materials Figure S6). Then the selected principal components were fed into a linear support vector machine (SVM) classifier (regularization parameter C = 1; logistic regression classifier was also tried, please see Supplementary Materials Figure S6) for training a classification model. We used the trained model to classify the unseen test data (dataset 1) and evaluated the classification performance of the model. Furthermore we investigated classification performances of the trained model on chronic medicated SSD, first episode medicated SSD and first episode unmedicated SSD separately in dataset 1.

### Statistical analysis

Binomial test (29) was used to estimate the statistical significance of the accuracy by determining whether these performances exceeded chance levels. In classification of SSD and HCs, the random classification model has p = 50% chance to predict the label correctly. Therefore, the cumulative probability of correctly predicting not more than k corrected labels in total n labels can be calculated from the cumulative binomial distribution function:

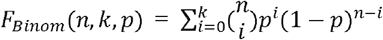

Therefore, the p value of accuracy is 1 – *F_Binom_*(*n,k,p*) The statistical significance level P < 0.05 was considered statistically significant. The binomial method has been used in previous study (30).

The classification model trained using chronic SSD from pooled dataset 2, dataset 3 and dataset 4 can identify chronic medicated SSD in dataset 1but cannot identify first episode SSD could reflect effect of illness duration and/or medication history or their interaction on FC (see Results section). Since there were very few chronic unmedicated SSD in this study (9 SSD), we could not use the two-factors analysis of variance, but used the following strategies to investigate the effect of illness duration or medication history on FC. We tested the effect of medication by comparing first episode medicated SSD with first episode unmedicated SSD to reduce illness duration confound. In addition, we tested the effect of illness duration by comparing chronic SSD with first episode medicated SSD to reduce medication confound. We used the network-based statistic (NBS) approach (31) to compare FC between groups. We took age, gender, education level, framewise displacement (FD) and illness duration as covariates to detect the effect of medication, while took age, gender, education level and FD as covariates to detect the effect of illness duration. Demographic and clinical information of chronic SSD, first episode medicated SSD and first episode unmedicated SSD were showed in Figure S2 (Supplementary Materials). The NBS is a nonparametric statistical method to deal with the multiple comparisons problem on a graph and control the family-wise error rate (FWER). We set the primary cluster-forming threshold to 3 (t statistics), and the corrected significance to 0.05 (two-tailed test) with 1000 times permutation.

### Correction of site and covariates

Site and covariates, e.g., gender and head motion may have a potential impact on our main findings. We additionally corrected the effect of site and covariates. Please see Supplementary Materials for details. Our main conclusions were not influenced.

## Results

### Data inspection of the four groups’ FC

As shown in Figure 2, the FC patterns and the distribution of FC of the four centers were very consistent. Correlations of connectivity of the four centers range from 0.89 to 0.95.

**Figure 2.**
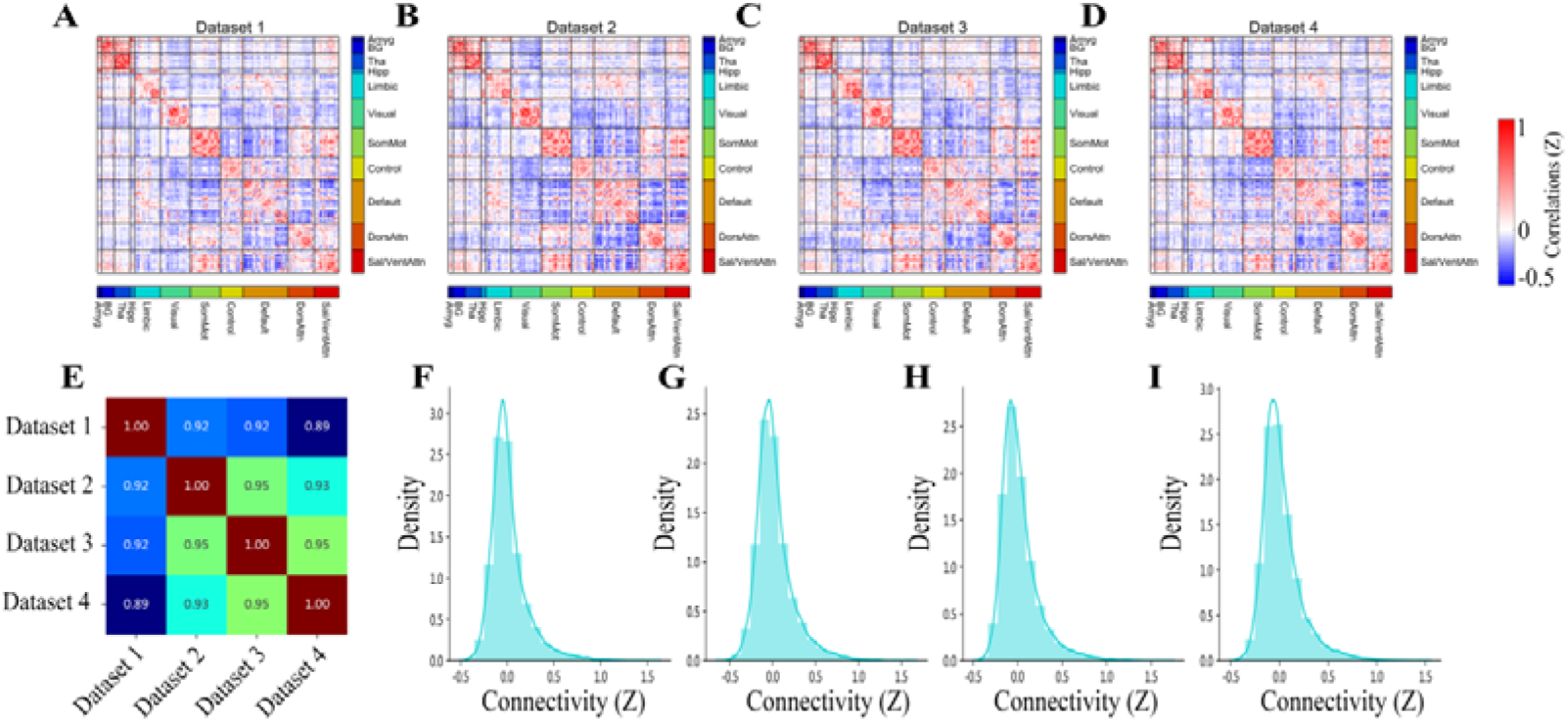
Data inspection of the four centers. (A-D) FC matrices of the four centers. (E) Correlations of connectivity of the four centers. (F-I) Distribution of connectivity of the four centers. Amyg = Amygdala; BG = Basal Ganglia; Tha = Thalamus; Hipp = Hippocampus; SomMot = Somatomotor; Control = Frontoparietal Control; Default = Default mode; DorsAttn = Dorsal Attention; Sal/VentAttn = Salience/Ventral attention.

### Classification performances

The classification model trained using chronic SSD from three sites (dataset 2, 3 and 4) classified SSD from HCs in another site (dataset 1) with 69% accuracy (P = 2.86e-13), 63% sensitivity, 75% specificity. Subgroup analysis indicated that this model can identify chronic medicated SSD in dataset 1 with 71% sensitivity (92 chronic medicated SSD; P = 4.63e-05), but cannot be generalized to first episode SSD, including first episode unmedicated SSD (n = 44; sensitivity = 48%, P = 0.68) and first episode medicated SSD (n = 74; sensitivity = 59%, P = 0.10). When we used 24 or 36 months as threshold to define the first episode and chronic SSD, the sensitivity of the first episode medicated SSD had slightly increased (Supplementary Materials Figure S5). When the difference of first episode and chronic stage was not considered, additional analyses also showed that this model cannot be generalized to unmedicated SSD, including unmedicated schizophreniform and unmedicated SZ (Figure S4 in Supplementary Materials).

For other three machine learning strategies: 5-fold cross-validation that pooled all datasets, five-fold cross-validation that only including first episode unmedicated SSD and leave-one-site-out cross-validation, all of the classification models achieved good performances when subgroup analyses were not used (please see the Supplementary Materials for details).

### Effect of illness duration or medication history on FC

We found first episode medicated SSD had different FC with first episode unmedicated SSD in a number of networks, e.g., the basal ganglia/striatum, visual and frontoparietal control networks (Figure 3). However, chronic SSD had nonsignificant different FC with first episode medicated SSD.

**Figure 3.**
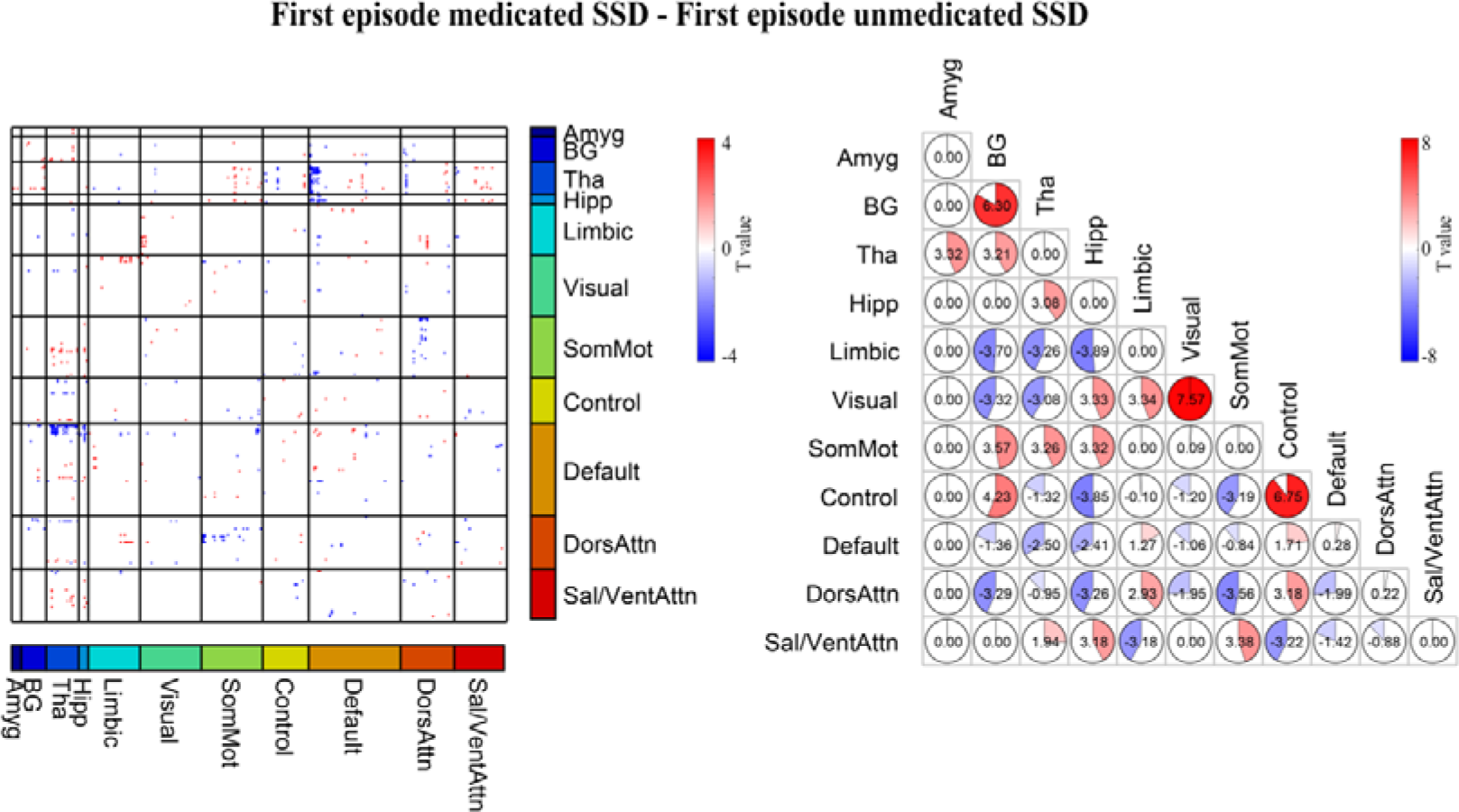
Differences of FC between first episode medicated SSD and first episode unmedicated SSD. (A) Differences were displayed as t-statistic values for each connectivity. (B) Differences were displayed as average t-statistic values within and between networks. Amyg = Amygdala; BG = Basal Ganglia; Tha = Thalamus; Hipp = Hippocampus; SomMot = Somatomotor; Control = Frontoparietal Control; Default = Default mode; DorsAttn = Dorsal Attention; Sal/VentAttn = Salience/Ventral attention; SSD = Schizophrenia Spectrum Disorder.

## Discussion

In the present study, we took FC as an example of neuroimaging-marker and using linear SVM as classifier to investigate whether or not the classification model trained using chronic SSD identified the trait biomarker of SSD that can be generalized to early stage SSD at the individual level by using a large multi-center samples. We found that although the classification model trained using chronic medicated SSD from dataset 2, 3 and 4 can identify chronic medicated SSD in dataset 1, but cannot be generalized to first episode SSD, including first episode unmedicated SSD and first episode medicated SSD. Univariable analysis indicated that medication usage had significant effect on FC, but illness duration had no significant effect on FC. Collectively, these findings suggest that the classification model trained using chronic medicated SSD may mainly find the pattern of chronic medication usage state, rather than identified the trait biomarker of SSD. Therefore, we should reconsider the current machine learning studies in chronic medicated SSD more cautiously in term of the clinical application, and highlight effect of medication usage on FC.

### Classification model trained using chronic SSD from dataset 2, 3 and 4 can identify chronic SSD in dataset 1

These findings were consistent with previous studies that found neuroimaging combined with machine learning can classify chronic medicated SSD (11, 13, 15, 19, 32–35). Besides, when subgroup analyses were not used, we also found functional connectivity combined with SVM can identify SSD with satisfied classification performances among three machine learning strategy: the 5-fold cross-validation that pooled all datasets, the leave-one-site-out cross-validation and the five-fold cross-validation that only including first episode unmedicated SSD (Supplementary Materials). These findings also in line with previous studies that found neuroimaging combined with machine learning can identify first episode SSD (36) or first episode unmedicated SSD (12, 37, 38).

### Classification model trained using chronic SSD from dataset 2, 3 and 4 can cannot be generalized to first episode SSD in dataset 1

An accurate identification of schizophrenia spectrum disorder (SSD) at early stage could potentially allow for treating SSD with appropriate intervention to potentially prevent future deterioration (25, 26, 39). If the classification model identified the trait biomarker of SSD, then we may expect it can diagnose SSD at the early stage of this disease. Unfortunately, our study found the classification model trained using chronic SSD can cannot be generalized to first episode SSD, especially the first episode unmedicated SSD. This finding supports and extends the idea of a previous study (21) that we should reconsider current evidence for the diagnostic value of machine learning and neuroimaging more cautiously. Beyond first episode SSD, additional analysis showed that the classification model also cannot be generalized to unmedicated SZ which had relatively longer duration than first episode SSD (Figure S4 in Supplementary Materials; note that there was an intersection between unmedicated SZ and first episode SSD). This finding further suggests that the failure of generalization cannot all be attributed to the short illness duration of first episode SSD in dataset 1.

### Medication usage had an significant effect on FC

We found medication usage had significant effect on FC, but disease duration had no significant effect on FC. This finding was an extension and supplement of a previous study (40). This previous study compared ultra high-risk subjects, first episode SSD and chronic SSD with HCs separately, and provide evidence for distinct patterns of functional-dysconnectivity across first episode and chronic SSD. As an extension, our study suggests that the distinct patterns of dysconnectivity across first episode and chronic SSD may be related to medication usage rather than illness duration. A recently study found illness duration had no effect on any cognitive domain when totally control the medication by using never-medicated individuals on the SSD (41). This finding partially supports the current finding. In addition, although we did not detect significant effect of illness duration on FC, classification model trained using chronic SSD also can cannot be generalized to first episode medicated SSD may suggest the interaction effect of medication usage and illness duration on FC.

### Limitation

First, we only used FC as feature to diagnose SSD, which may be a main concern. However, our machine learning pipeline using FC achieved good performance to identify chronic SSD as well as good performance using other strategies to identify first episode unmedicated SSD compared to previous study (Figure S1 in Supplementary Materials). These findings suggest that the failed generalization to first episode SSD was not due to the selection of feature or machine learning method. Besides, considering that a large number of published studies used function metrics to classify chronic SSD, it is may necessary to test the real clinical application value. Second, ideal statistical method to test the effect of medication usage or/and illness duration on FC is the two-factors analysis of variance. However, since there were very few cases of chronic unmedicated SSD, we were unable to use this method the two-factors analysis of variance.

### Conclusion

In conclusion, we found that the classification model trained using chronic medicated SSD can successfully identify chronic medicated SSD, but cannot be generalized to first episode SSD, especially the unmedicated SSD. Univariable analysis showed that medication usage had significant effect on FC, but disease duration had no significant effect on FC. These findings suggest that the classification model trained using chronic medicated SSD may mainly identified the pattern of chronic medication usage state, rather than the trait biomarker of SSD. We should reconsider the current machine learning studies in chronic medicated SSD more cautiously in term of the clinical application.

## Data Availability

The data used in this work can be requested from the corresponding author

## Acknowledgment

This work was supported by National Key R&D Program of China (Grant 2018YFC1311600 and 2016YFC1306900 to Yanqing Tang), Liaoning Revitalization Talents Program (Grant XLYC1808036 to Yanqing Tang), Science and Technology Plan Program of Liaoning Province (2015225018 to Yanqing Tang), National Science Fund for Distinguished Young Scholars (81725005 to Fei Wang), Liaoning Education Foundation (Pandeng Scholar to Fei Wang), Innovation Team Support Plan of Higher Education of Liaoning Province (LT2017007 to Fei Wang), Major Special Construction Plan of China Medical University (3110117059 and 3110118055 to Fei Wang).

## Conflict of interest

The authors declare no conflict of interest.

## Notes

### Competing Interest Statement

The authors have declared no competing interest.

### Author Declarations

Ethics Committee of the First Affiliated Hospital of China Medical University

